# Using e-vouchers to promote use of a family planning call center: A comparative interruptive time series analysis

**DOI:** 10.1101/2023.11.26.23298739

**Authors:** Dominique Meekers, Lynn Abu Turk, Olaniyi Olutola

**Affiliations:** Department of International Health and Sustainable Development, School of Public Health and Tropical Medicine, Tulane University, New Orleans, LA, USA; Sponsored Projects Administration, Tulane University, New Orleans, LA, USA; Department of Epidemiology and Medical Statistics, Faculty of Public Health, University of Ibadan, Ibadan, Nigeria

**Keywords:** family planning, call center, e-voucher campaign, impact evaluation, Nigeria

## Abstract

**Introduction:** This study examines the effectiveness of an e-voucher campaign to promote the use of a toll-free family planning call center (Honey&Banana), which is measured by the number of calls received by the call center.

**Methods:** A comparative interrupted time series design is used to measure the effect of the e-voucher campaign on the call volume. The study design presumes that without the e-voucher campaign, the intervention and control groups would have parallel trends in terms of the number of calls received. The difference between the actual and anticipated number of calls received is used to estimate the effect of the campaign.

**Results:** Before the e-voucher campaign, the monthly trend in the number of calls for family planning information in the intervention and control groups followed a similar pattern. As soon as the campaign was launched, the call center experienced a seven-fold increase in the number of family planning requests that were received from callers in the intervention zone, from under 400 calls per month to about 3,000 calls per month. No comparable increase occurred for the control zone. We estimate that the e-voucher campaign resulted in a net gain of over 12,000 requests for family planning information. When the campaign ended, the number of calls from the intervention zone decreases, but at least temporarily remained above the pre-intervention levels.

**Discussion:** Earlier studies demonstrated that e-voucher campaigns can be effective for increasing use of smoking cessation hotlines. This study expands the evidence base by demonstrating the effectiveness of e-vouchers for increasing demand for family planning information hotlines. Additional research is needed to assess the long-term effects of such e-voucher campaigns, and to determine the extent to which increased use of family planning hotlines translates into improved family planning outcomes.

## 1 Introduction

Voucher campaigns have been used extensively to promote a wide range of health services (Ali, Azmat, Hamza, Rahman, & Hameed, 2019; Arur et al., 2009; Azmat, Shaikh, Hameed, Mustafa, & Hussain, 2013; Bajracharya, Veasnakiry, Rathavy, & Bellows, 2016; B. Bellows, Bajracharya, Bulaya, & Inambwae, 2015; B. Bellows et al., 2016; Burke, Gold, Razafinirinasoa, & Mackay, 2017; Eva, Quinn, & Ngo, 2015; McConnell, Rothschild, Ettenger, Muigai, & Cohen, 2018). Typically, vouchers can be taken to an approved provider in exchange for health products or services. With respect to family planning programs, voucher campaigns have been identified as a proven High Impact Practice Enhancement (High Impact Practices in Family Planning (HIP), 2020; Riley, 2019). Although a considerable body of literature documents the effects of voucher programs on key family planning outcomes, there are large variations in how voucher programs have been implemented. Hence, more research is needed to share program experiences and document what works and what does not (Gorter, Grainger, Okal, & Bellows, 2012; Menotti & Farrell, 2016). Given that voucher programs are increasingly shifting from paper-based vouchers to electronic vouchers (so-called e-vouchers) (High Impact Practices in Family Planning (HIP), 2020; Lee & Adam, 2021; Riley, 2019), more evidence is also needed on the effectiveness of various ways of implementing e-vouchers programs. While there is evidence voucher programs are an effective way to reduce financial barriers to traditional health services, few studies have examined the effectiveness of vouchers for encouraging use of health services that are not well-known or new types of new health services (Gorter et al., 2012). That is also the case for e-voucher programs that promote the use of family planning hotlines or call centers.

Research on smoking cessation hotlines has consistently shown that offering incentives, such as free nicotine patches or gum, substantially increases call volumes to these quitlines (An, Schillo, Kavanaugh, Luxenberg, et al., 2006; An, Schillo, Kavanaugh, Lachter, et al., 2006; Anderson, Kirby, Tong, Kohatsu, & Zhu, 2018; Bauer, Carlin-Menter, Celestino, Hyland, & Cummings, 2006; Cummings et al., 2006; Hood-Medland et al., 2018; Momin et al., 2014; Tong et al., 2023; Vijayaraghavan et al., 2018). Although several organizations operate family planning call centers (Antillon, Webbe, & Husken, 2022; Corker, 2010; Kulathinal, Joseph, & Saavala, 2019; Yagnik et al., 2015), to the best of our knowledge, there are no studies that examine whether e-vouchers can be effective for increasing use of such family planning hotlines.

This paper uses a comparative interrupted time series design to estimate the effect of an e-voucher campaign to promote use of the *Honey&Banana* family planning call center in Nigeria. We measure impact of the *Honey&Banana* e-voucher campaign on the volume of calls received by the call center using an interrupted time series design. The study results will help the program make informed decisions about campaigns to promote use of the call center and helps build the evidence base about the potential for voucher campaigns to increase use of family planning hotlines.

## 2 Background

In 2017, DKT International Nigeria launched *The Honey & Banana Connect* (H&B) program, a multi-component platform that aimed to simplify contraceptive decision-making by providing confidential family planning information and services, with a focus on long-acting reversible contraceptive methods. The H&B platform consists of multiple components. The main component is a toll-free call center, which is supplemented by website, phone app, and various social media. The call center is linked to DKT nationwide network of family planning clinics. The call center’s toll-free number enables customers to confidentially obtain family planning information from a trained agent. Because Nigeria is linguistically very diverse, H&B ensures that the call center is able to provide information in five of Nigeria’s main languages (English, Hausa, Igbo, Yoruba, and Pidgin English). For callers who are interested in obtaining a contraceptive method, call center agents will help identify a nearby trained family planning provider from the affiliated clinic network and refer the caller to that location (DKT International Nigeria, 2021).

H&B conducts various activities that aim to promote the call center and increase the number of customers who take advantage of its free contraceptive information services. For example, from December 2021 through March 2022, H&B conducted a radio jingle campaign to promote the call center. Although that campaign increased the number of callers, it was not as successful as desired (Meekers, Olutola, & Turk, 2023b). Because sporadic small-scale e-voucher campaigns for free contraceptives had shown a lot of promise, H&B launched a large e-voucher campaign from June 2022 through November 2022.

Electronic vouchers, or e-vouchers are widely used to reduce financial obstacles to family planning (Ali et al., 2019; Arur et al., 2009; Azmat, Shaikh, et al., 2013; Bajracharya et al., 2016; B. Bellows et al., 2015; B. Bellows et al., 2016; Boddam-Whetham, Gul, Al-Kobati, & Gorter, 2016; Burke et al., 2017; Eva et al., 2015; High Impact Practices in Family Planning (HIP), 2020; McConnell et al., 2018; Riley, 2019). In addition to reducing the cost of family planning products and services, vouchers can facilitate access to private sector providers. Increased access to the private sector may in turn increase family planning use because of the perception that the private sector offers high quality, confidential services (High Impact Practices in Family Planning (HIP), 2020). There is growing evidence that voucher programs have the potential to improve family planning outcomes, such as contraceptive knowledge, contraceptive uptake and contraceptive equity, when used in conjunction with a larger family planning program (Ali et al., 2019; Atukunda et al., 2019; Azmat, Mustafa, et al., 2013; Burke et al., 2017).

Vouchers can also be a useful approach for promoting new types of services that people are not familiar with, or that they are hesitant to use. Voucher recipients are believed to consider it like a personal invitation, that then empowers them to use the services (Gorter et al., 2012; Grainger, Gorter, Al-Kobati, & Boddam-Whetham, 2017). For example, several organizations have used vouchers to increase use of smoking cessation helplines. Evidence shows that providing free nicotine replacement therapy (NRT), tends to substantially increase the number of people who call the hotlines (An, Schillo, Kavanaugh, Luxenberg, et al., 2006; An, Schillo, Kavanaugh, Lachter, et al., 2006; Anderson et al., 2018; Bauer et al., 2006; Cummings et al., 2006; Hood-Medland et al., 2018; Tong et al., 2023; Vijayaraghavan et al., 2018). For example, a study of the Minnesota QUITPLAN Helpline shows that offering free NRT increased the number of callers from 155 to 679 per month (An, Schillo, Kavanaugh, Lachter, et al., 2006). Similarly, a study of the California Smokers’ Helpline found that offering either free nicotine patches or a gift card was associated with a four-fold increase in the probability of calling the hotline, and offering both patches and a gift care increased the call probability by a factor of seven (Anderson et al., 2018). Although it is evident that voucher programs can drastically increase call volumes for smoking cessation hotlines, there is a lack of evidence on the potential effect of voucher campaigns on call volumes for family planning hotlines, such as the H&B call center.

The main aim of the H&B e-voucher campaign was to raise awareness of the H&B call center and to generate an increase in the number of customers to contact the call center to request family planning information and/or a referral to a family planning provider. The e-voucher campaign was geographically targeted to nine states. Eight of these states were selected because they are disadvantaged areas that are priority states for the call center’s main funding agency, the Bill and Melinda Gates Foundation (Bauchi, Gombe, Kaduna, Kano, Lagos, Nasarawa, Niger, and Sokoto). The campaign was extended to include Enugu state with funding from the Packard Foundation. E-vouchers were not offered in the remaining 27 states or in the Federal Capital Territory.

Because voucher sensitization and awareness activities are essential for the success of voucher programs (Chattopadhyay, Townsend, & RamaRao, 2015; Lee & Adam, 2021; Menotti & Farrell, 2016), the availability of e-vouchers for free contraceptives in the nine intervention states was advertised via several channels, including short ads on several radio stations in the target states, through Honey&Banana’s Facebook and Instagram pages, and through several Hausa social media influencers and blogs (e.g., Arewafamilyweddings, Diaryofanorthernwoman, Hausaroom, Hausafulanii, Kannywood, Northerners_blog, Northernhibiscus, and Northernmarriages).

Since the objective of our study is to estimate the net impact of the e-voucher campaign, it is important to consider that the number of incoming calls may have been affected by other confounding factors, such as other H&B promotional activities. For example, H&B continuously promotes the call center’s toll-free number through its website and social media, and the call center number is printed on most DKT’s contraceptive products. Since such promotional activities were conducted in both the intervention and control groups, we are confident that they did not disproportionately affect the e-voucher intervention states.

In theory, activities by other family planning organizations (including other non-governmental organizations and the government) could also increase the demand for family planning information, which may increase the number of incoming calls. However, most of these programs are nationwide, and should therefore affect the e-voucher intervention and control states similarly. Furthermore, we are not aware of major program activities that coincided with the intervention period of the H&B e-voucher campaign. Therefore, family planning programs by the government or other NGOS are unlikely to be an important confounder.

## 3 Data and methods

### 3.1.1 Study Design

We estimate the effect of the e-voucher campaign on the number of calls received by the H&B family planning call center using a comparative interrupted time series design. In public health, interrupted times series (ITS) are commonly used to measure the effect of policies and programs on health outcomes, particularly when it is not feasible or desirable to conduct a randomized controlled trial (Booth, Allen, Bray Jenkyn, Li, & Shariff, 2018; Carson et al., 2017; Cheng et al., 2016; Hategeka, Ruton, Karamouzian, Lynd, & Law, 2020; Hudson, Fielding, & Ramsay, 2019; Humphreys, Gasparrini, & Wiebe, 2017; Jandoc, Burden, Mamdani, Levesque, & Cadarette, 2015; Ramsay, Matowe, Grilli, Grimshaw, & Thomas, 2003; Siedner et al., 2020; Turner et al., 2020). ITS study designs are most appropriate for evaluating interventions that have a clearly defined starting point, and for which several pre- and post-intervention data points are available. As such, ITS can be suitable for evaluating the effect of family planning interventions – including e-vouchers -on routinely collected family planning outcome indicators (Fuseini, Jarvis, Hindin, Issah, & Ankomah, 2022; Ma, Cecil, Bottle, French, & Saxena, 2020).

The simplest form of ITS design examines the trend in a key outcome measure for a single group, without a comparison group. It is expected that the effect of the intervention should be clearly visible in the trend line. Specifically, if the intervention is effect, the level and/or slope of the outcomes measure should change noticeably change soon after the start of the intervention (Bernal, Cummins, & Gasparrini, 2017). Although an interruption in the trend line is indicative of a successful intervention, it could also be caused by other confounding factors (e.g., by other interventions). This concern can be addressed by using a comparative interrupted time series design (CITS). The CITS adds a comparison group that was not exposed to the intervention, but that is presumed to have experienced the same confounding factors (Bernal, Cummins, & Gasparini, 2018; Bottomley, Scott, & Isham, 2019). When randomized controlled trials are not feasible, CITS are considered among the strongest study design for measuring program impact (Fretheim, Soumerai, Zhang, Oxman, & Ross-Degnan, 2013; Fretheim et al., 2015; St.Clair, Cook, & Hallberg, 2014; St.Clair, Hallberg, & Cook, 2016).

Because the H&B e-voucher campaign was limited to nine specific states, we measure the effect of the campaign using an intent-to-treat approach that compares these intervention states with a control group that consists of regions where the e-vouchers were not offered. If the e-voucher campaign was effective, then we would expect to see a notable increase in the number of calls received from the intervention states, but not elsewhere. Because an e-voucher code can only be obtained by contact the call center, we anticipated that the start of campaign activities to inform the public about the availability of the e-vouchers (e.g., social media posts, radio hype) would immediately lead to an increased in the volume of calls received. Hence, we anticipate seeing an intervention effect – as evidenced by an interruption in the times of the number of calls received -- from June 2022 onward.

We estimate the impact of the e-voucher campaign using the parallel trends assumption. Specifically, we assume that in absence of e-voucher campaign, the trend in the number of calls received from the intervention states would have the same shape as the trend in the number of calls received from the control regions. In other words, we argue that the trend in the control region reflects the secular trend in the number of calls that the call center would have received in absence of the e-voucher campaign.

The parallel trends assumption is premised on the belief that any confounding factors that affect the number of calls would affect both groups similarly (Bernal et al., 2018; Bottomley et al., 2019; Degli Esposti et al., 2021). To confirm this, we graphed the pre-intervention trends in the intervention and control locations. The graphical presentation of the CITS results is easy to interpret and can send a convincing message about program impact, even prior to statistical analyses to measure the magnitude of the intervention impact (Jandoc et al., 2015; Penfold & Zhang, 2013; Wagner, Soumerai, Zhang, & Ross-Degnan, 2002). Because some segments of the population may be more responsive to the intervention or to confounding interventions, it is recommended to check whether the intervention and control group have similar characteristics (Bernal et al., 2018). Therefore, we compared the characteristics of women living in the intervention and control areas using recent survey data (National Population Commission (NPC)[Nigeria] & ICF, 2019).

After confirming that intervention and control groups are roughly comparable and had similar pre-intervention trends in the number of incoming calls, we estimated the projected number of calls that would have happened in absence of the e-voucher campaign. Specifically, we assume that without the intervention, the trends in the number of incoming calls from intervention region would have had the exact same shape as the trend in the control group (i.e., a parallel trends), albeit with a different starting point. We estimate the effect of the e-voucher campaign on the number of incoming calls by calculating the difference between the actual and projected monthly number of calls received after the intervention started.

### 3.1.2 Data source

Our interrupted times series analysis is based on routine data on the number of calls received by the call center, which are available in a public data repository (Olutola, Inyang, & Nwaogbo, 2023). During the two-year period from January 2021 to December 2022 the call center received a total of 33,552 calls. The published call volumes exclude calls that were not related to family planning (e.g., wrong number, cranks calls, calls on topics outside the scope of the call center). The available data consist of the monthly number of incoming calls received by the call center, disaggregated by state of origin of the call.

For the purposes of this study, the intervention area was defined as the nine states where the e-voucher campaign was implemented (Bauchi, Enugu, Gombe, Kaduna, Kano, Lagos, Nasarawa, Niger, and Sokoto). The control area consists of the remaining 27 states and the Federal Capital Territory (FCT).

We use an intent-to-treat analysis in which all callers who reside in the intervention states are considered to have been exposed to the e-voucher intervention, irrespective of whether they were exposed to any of the activities to increase awareness of the e-voucher campaign.

### 3.1.3 Study limitations

Although use of a CITS design with a control group can provide strong evidence of program impact, it is impossible to know exactly how many incoming calls would have originated from the intervention states without the e-voucher campaign. Based on our examination of the comparability of the characteristics of the intervention and comparison group, and the similarity in the pre-intervention trends in the number of number of incoming calls, we are confident that the parallel trends assumption is reasonable.

There is a chance that our results may have been affected by contamination (i.e., some members of the control group may have been exposed to the intervention). In theory, it is possible that some residents in the control group may have become aware of the e-voucher campaign and may have contacted the call center in the hopes of obtaining a voucher code. Because the campaign materials clearly mentioned which states participated in the e-voucher campaign, we believe such events to be rare. If a caller from a non-participating state did request a voucher, he or she would have been advised that such vouchers can only be redeemed at family planning providers in the participating states. Hence, we believe the risk of contamination is minimal.

Because our data are limited to the period through December 2022, we are unable to access any potential longer term effects of the e-voucher campaign on the call volume. Although a longer data series would be preferable, the call center re-located in January 2023 and was assigned a different toll-free number. Because this affect the call volumes in early 2023, it is not possible to assess any longer term effects of the e-voucher campaign.

## 4 Results

The strength of the CITS study design relies on the assumption that the intervention and control groups have similar exposure to confounding factors that may affect the number of incoming calls, and that the comparison group is a good counterfactual. To check whether that is a reasonable assumption, we verified 1) whether two groups have comparable the socio-demographic characteristics, and 2) whether the pre-intervention trends in the two groups followed a similar pattern.

### 4.1.1 Characteristics of the intervention and control groups

Because it is desirable for the intervention and control group have similar characteristic (Bernal et al., 2018), we checked for covariate balance between two groups using 2018 Nigeria Demographic and Health Survey. Because the call center is known to have predominantly female customers, our analysis of the DHS data is limited to women of reproductive age. Table 1 in Supplementary File 1 indicates that women living in the intervention states and control states are fairly similar in terms of the level urbanization, educational level, wealth status, and phone ownership. However, because several of the intervention states are located in the North, women in the intervention area are much more likely than those in the control areas to be Hausa (46% vs 22%) and Islamic (71% vs. 45%).

**Table 1:** Estimation of the number of family planning calls gained by the e-voucher campaign (July - November 2022)

|  | Control<br>group | Intervention group |  | Intervention<br>group |
| --- | --- | --- | --- | --- |
|  | Observed | Observed | Predicted* | Gained |
| 06/22 | 908 | 1,637 | 665 | 972 |
| 07/22 | 888 | 2,914 | 645 | 2,269 |
| 08/22 | 892 | 2,839 | 649 | 2,190 |
| 09/22 | 785 | 3,285 | 542 | 2,743 |
| 10/22 | 679 | 2,921 | 436 | 2,485 |
| 11/22 | 664 | 1,470 | 421 | 1049 |
| 12/22 | 477 | 703 | 234 | 469 |
| Total | 5,293 | 4,715 | 3,592 | 12,177 |

\* CITS comparative interrupted time series results.

The two groups have a fairly similar distribution by age and marital status, and number of children ever born. Moreover, the percentage of women in the intervention and control areas who know at least one modern contraceptive method is nearly identical (90% vs 93%), as is the percentage who currently use a modern method (11% each). In other words, despite the clear ethnic and religious differences between the two areas, they are very comparable in terms of demographics and family planning indicators.

However, it is noteworthy that the intervention states have a smaller total population size than the comparison states (67 vs. 139 million), which could affect the number of calls originating from each region (National Bureau of Statistics [Nigeria], 2021).

### 4.1.2 Comparison of the pre-intervention trends

The CITS study design on the assumption that the intervention and control group would have experienced parallel trends in absence of the intervention. Hence, we investigated whether there were comparable pre-intervention trends in the number of family planning related calls between the intervention and control groups. Figure 1 displays the trends in the quantity of requests for family planning information from callers in the states where the e-voucher campaign was implemented and those where it was not, with the intervention period marked in green.

**Figure 1:**
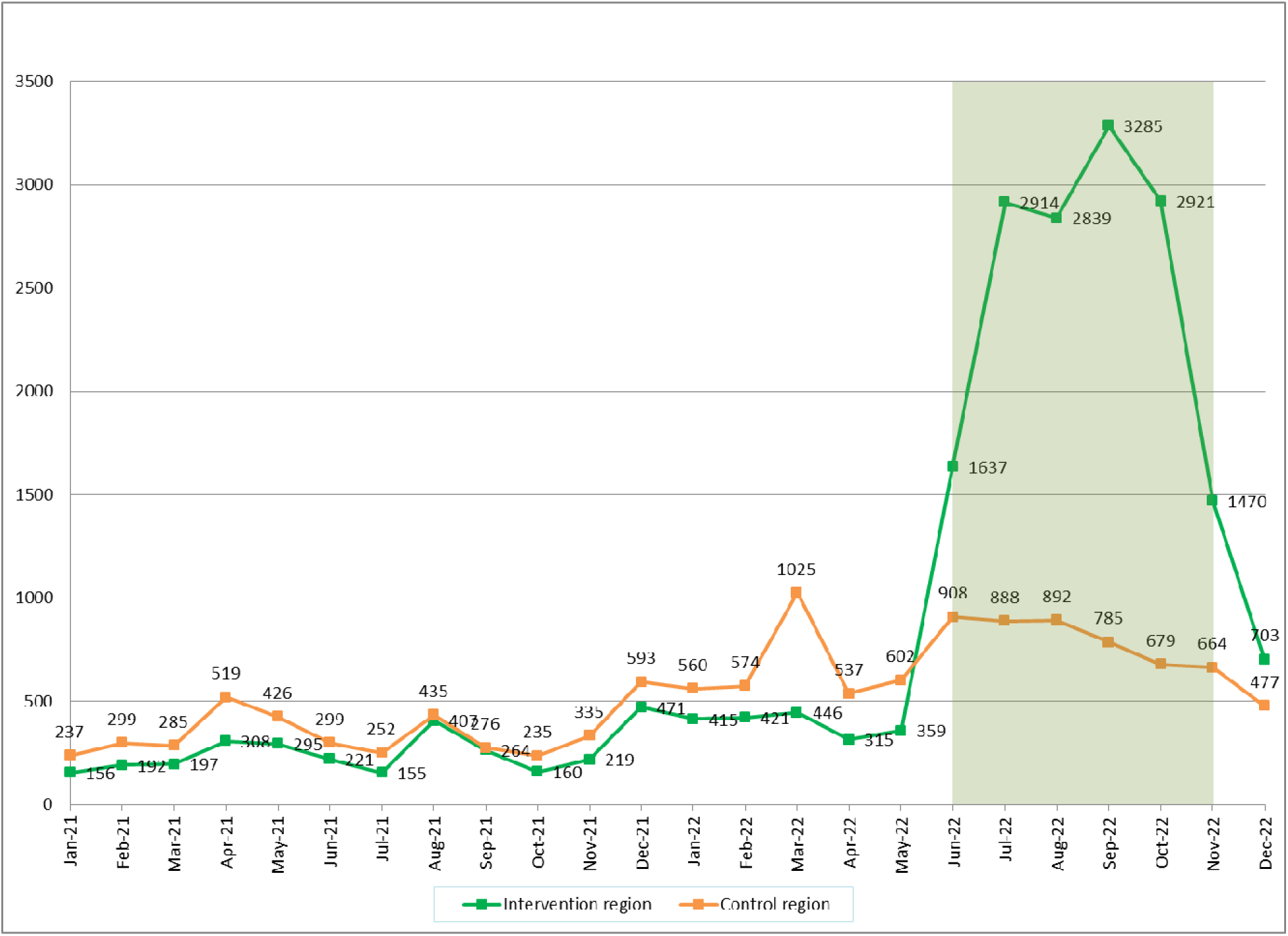
Number of requests for family planning information received, by intervention area

Figure 1 displays the trends in the volume of requests for family planning information from callers in the states where the e-voucher campaign was implemented and those where it was not, with the intervention period marked in green. Before the intervention (January 2021-May 2022), the number of calls that originated from the control region was consistently higher than that from the intervention area. This was expected, as the control region has a substantially larger population size than the intervention regions (National Bureau of Statistics [Nigeria], 2021). The trends in both regions had a nearly identical pattern. The small peak in the number of calls in April 2021 coincides with a small H&B campaign for Sayana Press injectable contraceptives, while the peak in August 2021 corresponds with promotional activities to celebrate the third anniversary of the call center (News Wings, 2021). The increased volume of calls between December 2021 and March 2022 reflects the effect of a radio campaign to promote the call center (Meekers, Olutola, & Turk, 2023a; Meekers et al., 2023b). The radio campaign was broadcast in twelve states, eight of which were located in the control group for the e-voucher campaign. In March 2022, Honey&Banana also conducted an HIV self-testing campaign at post-secondary educational institutions that encouraged customers to contact the call center with questions or concerns. Except for March 2022, the similarity of the pre-intervention trends in the intervention and control regions indicates that the study design controlled for confounding factors that affected both areas, such as nationwide program activities.

Calculation of the pre-intervention trend lines for the two regions confirms that the trend in the control states has a higher intercept, and a larger slope. The linear trend in the intervention region was 13.86 169.43 compared to 26.189 204.83 for the control region. Thus, the control area started with a higher number of calls than the intervention area. The steeper slope in the control area is likely largely due to the peak in the number of calls in March 2022.

### 4.1.3 Comparison of the post-intervention trends

As shown in Figure 1, as soon as the e-voucher campaign started in June 2022, the number of the number of incoming calls that originated from the intervention states increased dramatically while the number of calls from the control states increased only modestly. During the entire campaign period (June-November) calls from intervention states substantially outnumbered calls from control states, which provides compelling evidence that the e-voucher campaign increased use of the call center.

The number of calls from intervention states increased sharply during period from June through September -- the most intense campaign months -- and then gradually declined through December. During this same time period, incoming call from the control states increase in June, but steadily decreased afterward. Estimation of the trend line confirms this negative slope in the number of calls from the control region (69.786 1035.3).

The slightly elevated number of calls from the control states during the period from June through August suggests that they may have been some minor treatment contamination. Although the e-vouchers cannot be redeemed at family planning providers in the control, we cannot rule out that some customers may still have called. If so, then the parallel trends assumption will slightly underestimate the true effect of the e-voucher campaign.

### 4.1.4 Estimation of the campaign effect

To calculate the projected number of incoming calls from the intervention states that the call center would have received in absence of the e-voucher campaign, we assume that the trend in the non-intervention states is a good estimate of the underlying secular trend. In other words, we argue that in absence of the e-voucher campaign the number of incoming calls from intervention and non-intervention states would exhibit a parallel trend. In other words, had the e-voucher campaign not be implemented, the trend in the intervention region would have had the exact same shape as in the control group, although with a lower starting level. Prior to the start of e-voucher campaign, the call center consistently received fewer calls from the ten intervention states than from the control states (see Figure 2). In the final month before the e-voucher campaign, May 2022, the call center received 359 calls that originated from an intervention states, compared to 602 from one of the control states. The blue line in Figure 2 shows the predictor number of calls from an intervention states (in absence of the e-vouchers) using the parallel trends assumption.

**Figure 2:**
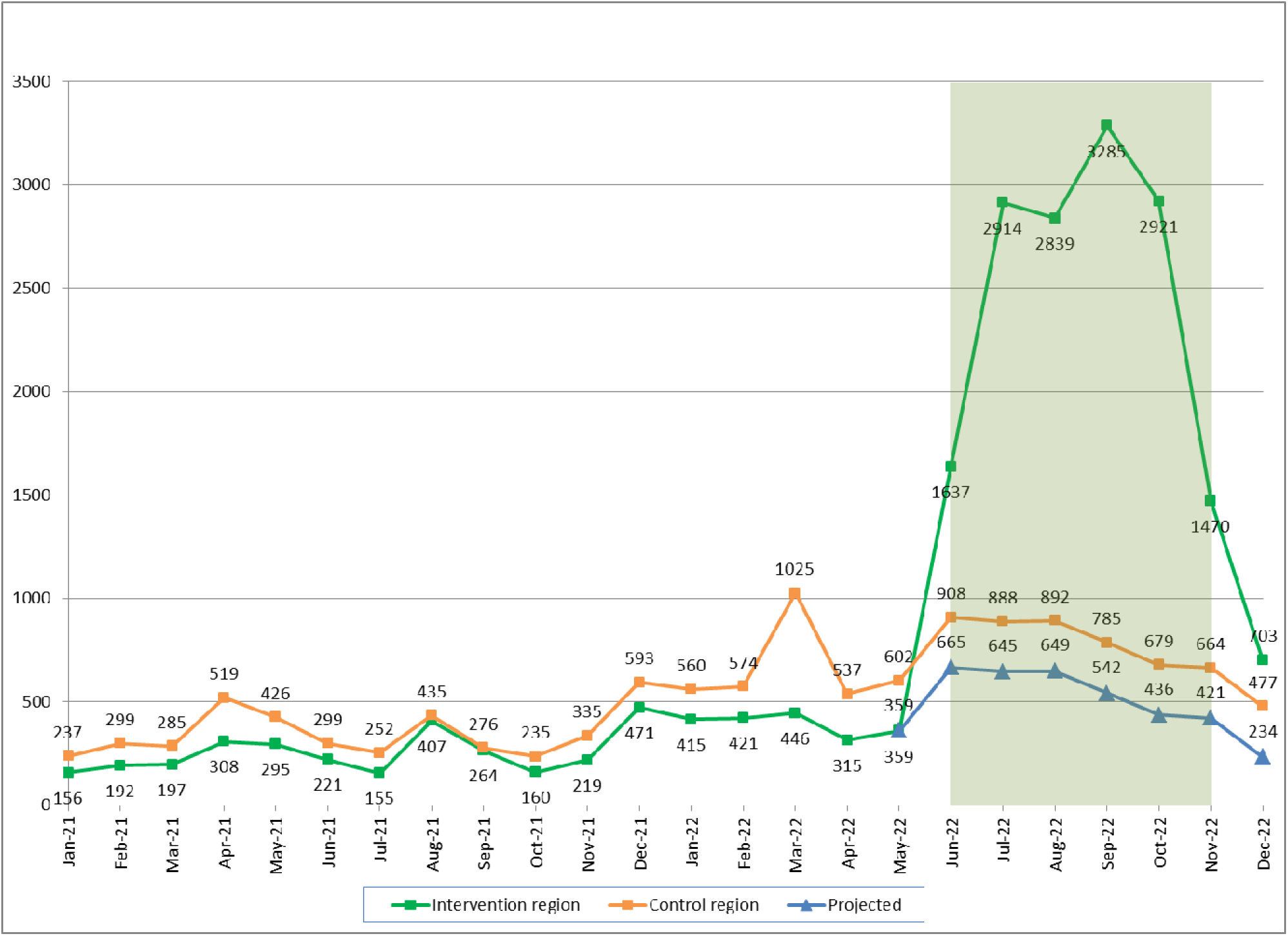
Actual and projected number of requests for family planning information, by intervention area

Comparison of the projected and observed number of calls from the intervention region (represented by the green and blue lines in Figure 2), illustrates the impact of the e-voucher campaign. The results indicate that the e-voucher campaign resulted in a drastic increase in the number of incoming calls. Following the start of the campaign, the actual number of incoming calls that originated from intervention states immediately increased from 359 in May to 1,637 in June. This drastic increased continued through September, when it reached a high of 3,285 calls. As the e-voucher campaign started to wind down, the observed number of calls decreased to 1,470 in November. Without the e-voucher campaign, the number of calls is projected to have increased only to 665 in June, and to have declined noticeably from September onward.

It is noteworthy that for December 2022, when the e-voucher campaign had already completely ended, the observed number of calls from the intervention states remained substantially higher than the number from the control states (703 vs. 477). This suggests that the e-voucher campaign could potentially have some longer-term effect on the number of incoming calls, for example by increasing awareness of the call center through word of mouth.

To quantify the increase in the total number of calls received as a result of the e-voucher campaign, we calculate the difference between the actual number of calls received and the projected numbers. Table 1 shows the actual and predicted number of incoming calls for the period from June 2022 thru December 2022, and the estimated number of calls gained due to the e-voucher campaign. During the first month (June 2002), the campaign generated an estimated 972 additional calls. As the campaign ramped up, the number of additional calls received roughly between 2,200 and 2,500 per month through October 2022. As the campaign wound down, the number of gained calls reduced to 1,049 in November. Although the e-voucher campaign officially ended in November 2022, an additional 469 calls were generated in December 2022. We are unable to discern whether this reflects callers who hoped to get an e-voucher past the deadline, or whether the e-voucher campaign increased demand for family planning information. In total, we estimate that the e-voucher campaign generated an additional 12,177 calls during this 7-month period. Considering that only 9,576 calls were received during the 12 months period to the e-voucher campaign, it is evident that the e-voucher campaign dramatically increased the number of incoming calls.

## 5 Discussion

Voucher campaigns have been recognized as an important high impact improvement for family planning programs and have been implemented extensively to reduce financial barriers to access contraceptive products and services. However, the implementation of these programs has varied considerably and more evidence about the effectiveness of different voucher strategies is needed. Evidence about the effectiveness of vouchers to promote new types of health services --or services that are relatively unknown-- is particularly scarce.

Taking advantage of the rapid increase in phone access, family planning programs in several countries have started to use call centers to increase access to quality sexual and reproductive health information (Antillon et al., 2022; Corker, 2010; Kulathinal et al., 2019). In Nigeria, DKT’s Honey&Banana call center provides confidential family planning information and counseling, free of charge. The toll-free number for the call center is posted on the project’s website and social media pages and is printed on the packaging for DKT’s contraceptive products, making it easily accessible. DKT has also been testing different approaches to further increase use of the H&B call center, such as the inclusion of promotional ads in the popular MTV Shuga Naija television series, and a radio jingle campaign to promote the call center. While research shows that the radio jingle campaign helped increase the number of H&B call center users, the increase was fairly modest. By contrast, a very small e-voucher campaign for free contraceptives conducted as part of the celebrations of the 3 ^rd^ anniversary of the call center generated considerable interest. That finding was the impetus for DKT to implement a large-scale e-voucher campaign to increase use of the H&B call center.

There is substantial evidence that voucher programs that aimed to increase the number of callers who use smoking cessation hotlines have been very successful (An, Schillo, Kavanaugh, Lachter, et al., 2006; Anderson et al., 2018; Bauer et al., 2006; Hood-Medland et al., 2018). However, we have not found any rigorous studies that examine the effectiveness of vouchers for increasing use of family planning hotlines. Our study contributes to the current state of knowledge by assessing the effect of an e-voucher campaign that aimed to increase the number of callers who used the H&B call center. Because the e-voucher campaign targeted only nine states, we were able to use a comparative interruptive time series design to measure the effect of the campaign. Comparative interrupted time series are among the recommended study designs for demonstrating the effectiveness of voucher programs (N. M. Bellows, Bellows, & Warren, 2011).

The results of our study provide convincing evidence that the e-voucher campaign resulted in a dramatic increase in the number of incoming requests for family planning information received by the H&B call center. Following the start of the campaign, the number of incoming calls from the callers located in the intervention zone increased from less than 400 calls to about 3,000 calls per month, approximately a seven-fold increase. This roughly seven-fold increase in calls from the intervention zone contrasts with the pattern for the control states, where no such increase was observed. We estimate that the e-voucher campaign resulted in a gain of at least 12,000 additional requests for family planning information. Because controlled interrupted time series are a strong study design when used in conjunction with a comparable control group, these results provide the first solid evidence that e-voucher campaign are not only effective for increasing use of smoking cessation hotlines (An, Schillo, Kavanaugh, Lachter, et al., 2006; Anderson et al., 2018; Bauer et al., 2006; Hood-Medland et al., 2018), but also an important tool for increasing use of family planning hotlines. In addition, it confirms that the e-voucher program can be valuable for increasing use of health services that are not yet well-known in the target population (Gorter et al., 2012; Menotti & Farrell, 2016).

Despite the clear impact of the H&B e-voucher campaign, our study also shows that the number of incoming calls from the intervention zones declined sharply as the campaign reached its end. Although the number of incoming calls in the month after the campaign (December 2022) remained substantially higher than the pre-campaign volume of calls, we cannot discern whether the campaign will have a long-term effect on call volumes. The main reason for our inability to do so is that the management of the H&B call center changed in January 2022, which resulted in a temporary closure of the call center (to allow for its physical relocation), and a change in the call center’s phone number from 55059 to 7790. Because of these disruptions, and various activities to promote the new phone number, subsequent data points are not comparable with the data presented in this paper. However, we concur with other authors (Azmat, Ali, & Rahman, 2023; B. Bellows et al., 2015; B. Bellows et al., 2016) that it is important for future studies to try and measure the longer-term effects, after the campaign has ended. In addition, it will be important for future studies to generate evidence about the extent to which increased of family planning hotlines translates into improved family planning outcomes, such as referrals to a family planning provider, adoption of modern contraceptive methods switching to more effective methods, and method continuation.

## Data Availability

All data produced are available online at

https://doi.org/10.7910/DVN/OMVLLB

https://dhsprogram.com

## Author Contributions

Meekers, D: Conceptualization, Formal Analysis, Writing – Original draft preparation; Abu Turk, L: Writing – Original draft preparation; Writing – Review & Editing; Investigation; Olutola, O: Data Curation; Writing – Review & Editing. All authors contributed to the critical revision of the manuscript and approved the final version.

## Acknowledgments

This research was conducted as part of the Honey & Banana Connect project, which is implemented by DKT Nigeria. The authors are grateful to Precious Nwaogbo, Vivian Obozekhai, and Edim Oluoma for providing information about the e-voucher campaign.

## Data Availability Statement

All call center data analyzed for this study are contained within the article. The raw data are available on Harvard Dataverse: Monthly number of phone calls received by the Honey&Banana Connect family planning call center in Nigeria (January 2021 – December 2022) https://doi.org/10.7910/DVN/OMVLLB (Olutola et al., 2023). The Nigeria DHS data are publicly available from https://dhsprogram.com.

## Ethics

Tulane University’s Human Research Protection Office determined that this study is not human subjects research and does not require IRB review and approval (Ref #2023-590).

---

**Supplementary Table 1.** Characteristics of women aged 15-49 in the intervention and control states.

|  | Intervention |  | Control |  |
| --- | --- | --- | --- | --- |
|  | % | No. | % | No. |
| **Type of place of residence** |  |  |  |  |
| Rural | 50.5 | 7,035 | 56.0 | 15,623 |
| Urban | 49.5 | 6,896 | 44.0 | 12,266 |
| **Level of education** |  |  |  |  |
| None | 43.4 | 6,053 | 30.7 | 8,550 |
| Primary | 12.1 | 1,680 | 15.6 | 4,358 |
| Secondary+ | 44.5 | 6,198 | 53.7 | 14,982 |
| **Wealth quintile** |  |  |  |  |
| Poorest | 17.2 | 2,398 | 17.3 | 4,825 |
| 2nd poorest | 21.0 | 2,927 | 18.3 | 5,118 |
| Middle | 17.5 | 2,440 | 20.7 | 5,767 |
| 2nd wealthiest | 18.0 | 2,505 | 23.3 | 6,485 |
| Wealthiest | 26.3 | 3,661 | 20.4 | 5,695 |
| **Owns mobile phone** |  |  |  |  |
| No | 49.1 | 6,843 | 42.5 | 11,845 |
| Yes | 50.9 | 7,089 | 57.5 | 16,045 |
| **Ethnic group** |  |  |  |  |
| Yoruba | 14.9 | 2,078 | 15.6 | 4,340 |
| Igbo | 10.7 | 1,488 | 17.7 | 4,931 |
| Hausa | 45.5 | 6,334 | 21.9 | 6,112 |
| Other | 28.9 | 4,031 | 44.8 | 12,506 |
| **Islamic** |  |  |  |  |
| No | 28.9 | 4,033 | 55.3 | 15,416 |
| Yes | 71.1 | 9,899 | 44.7 | 12,473 |
| **Age group** |  |  |  |  |
| 15-24 | 37.1 | 5,173 | 36.3 | 10,111 |
| 35-34 | 32.8 | 4,575 | 31.8 | 8,857 |
| 35-49=3 | 30.0 | 4,183 | 32.0 | 8,921 |
| **Currently married or cohabiting** |  |  |  |  |
| No | 27.1 | 3,780 | 32.1 | 8,951 |
| Yes | 72.9 | 10,151 | 67.9 | 18,938 |
| **Children ever born** |  |  |  |  |
| <3 | 49.8 | 6,940 | 52.3 | 14,584 |
| 3 or 4 | 19.8 | 2,754 | 20.3 | 5,653 |
| 5+ | 30.4 | 4,237 | 27.4 | 7,652 |
| **Knows at least one modern contraceptive method** |  |  |  |  |
| No | 10.1 | 1,410 | 6.8 | 1,890 |
| Yes | 89.9 | 12,522 | 93.2 | 26,000 |
| **Current family planning use** |  |  |  |  |
| None/folkloric | 86.7 | 12,085 | 86.0 | 23,988 |
| Traditional | 2.8 | 389 | 3.5 | 963 |
| Modern | 10.5 | 1,458 | 10.5 | 2,939 |
| Total | 100.0 | 13,932 | 100.0 | 27,889 |

Source: 2018 Nigeria Demographic and Health Survey, women aged 15-49 only. Weighted data.

